# Assessing the Reliability of a Controllable Sound Source–Driven Bowel Sound Monitoring Device in Physiological Tissue Acoustic Environments

**DOI:** 10.64898/2026.06.03.26354788

**Authors:** Zhao Jingxuan, Zhao Zhizhuang, Huang Xianya, Li Yuxuan, Wu Junling, Peng Shuhuang, Wang Shufang, Sun Gang, Luan Zhe

## Abstract

**Objective:** To verify the reliability of a self-developed bowel sound monitoring device under real biological tissue acoustic propagation conditions using a controllable sound source, and to establish quantitative evidence for its translational applicability.

**Methods:** Freshly euthanized six-month-old Bama miniature pigs were used as an experimental model. A high-fidelity Bluetooth audio playback device was implanted into the abdominal cavity to deliver manually annotated bowel sound recordings as controllable acoustic stimuli. A self-developed bowel sound monitoring device was fixed on the abdominal surface for continuous signal acquisition. Playback timestamps were defined as the ground truth, and event-level matching was performed within a predefined temporal tolerance window. Four performance indicators were evaluated: (1) bowel sound acquisition and energy amplification, (2) event matching accuracy, (3) acoustic feature consistency, and (4) subjective agreement assessed by blinded auscultation from gastroenterologists with different levels of clinical experience.

**Results:** The monitoring device exhibited stable detection capability and effectively covered the full spectral range of the original signals. It significantly enhanced bowel sound energy while preserving temporal and spectral characteristics, demonstrating high consistency in time- and frequency-domain features. Blinded clinician assessments showed a subjective agreement rate of 88.9% between original and surface-recorded bowel sound events.

**Conclusions:** Under real tissue acoustic propagation conditions, the self-developed bowel sound monitoring device reliably captures bowel sound events with high temporal accuracy, acoustic fidelity, and clinical perceptual consistency. This controllable sound source–based validation provides robust technical evidence for subsequent in vivo studies and clinical translation, supporting the development of objective and continuous gastrointestinal function monitoring.

---

Objective assessment of gastrointestinal functional status is essential for clinical decision-making and perioperative management [1]. Bowel sounds are acoustic signals generated during gastrointestinal peristalsis and intraluminal content propulsion, and can partially reflect changes in gastrointestinal motility. They have therefore been widely used in clinical physical examination and basic research [2]. However, conventional bowel sound assessment relies mainly on manual auscultation, which is subjective, intermittent, and difficult to quantify, thereby limiting its application in precision management and research analysis.

With advances in medical engineering and digital health technologies, sensor-based bowel sound monitoring and signal-processing approaches have attracted increasing attention [3]. Previous studies have explored the use of electronic stethoscopes, wearable sensors, and intelligent algorithms to acquire and analyse bowel sounds, with the aim of enabling continuous and objective assessment of gastrointestinal function [4]. Nevertheless, bowel sounds are typically sparse, short-lived, acoustically complex, and vulnerable to environmental noise, placing high demands on the acquisition performance and stability of monitoring devices [5]. Current studies have largely focused on signal processing and algorithm development, whereas the reliability of bowel sound monitoring hardware under realistic biological tissue propagation conditions remains insufficiently validated [6].

A key challenge in bowel sound research is the lack of a clearly defined and controllable reference for “true” bowel sound events [7]. In vivo, both the timing and acoustic morphology of bowel sounds are difficult to control precisely, and manual annotation inevitably introduces observer-dependent bias, increasing uncertainty in device performance evaluation [8]. This limitation has hindered the translation of bowel sound monitoring technologies from experimental research to clinical application. Therefore, before implementing complex algorithms or clinical studies, the fundamental acquisition capability of bowel sound monitoring devices should be independently validated from an engineering and methodological perspective.

In this study, we introduced a controlled acoustic source as a methodological strategy to validate the reliability of a bowel sound monitoring device under realistic biological tissue acoustic propagation conditions. Manually annotated bowel sound recordings were played within the abdominal cavity of freshly euthanized Bama miniature pigs, thereby establishing a defined acoustic propagation model while preserving the abdominal cavity–abdominal wall–body surface transmission pathway. This design minimized physiological uncertainty without aiming to reproduce active intestinal physiology. Instead, it focused specifically on the ability of the device to acquire and identify bowel sound events after propagation through real biological tissue.

We evaluated a self-developed bowel sound monitoring device, Bowelsoundset301, with respect to event detection capability, temporal localization accuracy, and resistance to false-positive detection. The findings are intended to provide an engineering basis for subsequent in vivo animal experiments, clinical validation, and algorithm development, and to support the standardization and clinical translation of bowel sound monitoring technologies.

## 1. Materials and Methods

### 1.1 Experimental animals

Freshly euthanized Bama miniature pigs were used in this study as an ex vivo biological tissue model. The animals were provided by the Large Animal Center of the Institute of Zoology, Chinese Academy of Sciences, and were used exclusively for scientific research. The selected animal was 6 months old and weighed 15.5 kg. All experimental procedures were completed within 6 h after euthanasia to preserve, as far as possible, the acoustic propagation properties of the abdominal cavity and abdominal wall tissues and to minimize the influence of post-mortem tissue changes on the results.

No additional interventions were performed on the animal during the experiment, and no live-animal procedures were involved. All procedures were conducted in accordance with relevant laboratory biosafety requirements and animal research ethics guidelines.

### 1.2 Methods

#### 1.2.1 Experimental device and system configuration

A self-developed bowel sound monitoring device was used as the body-surface acoustic signal acquisition system in this study. The device, protected by a Chinese invention patent (Patent No. ZL 2026 1 0178256.4), was designed for continuous monitoring and recording of bowel sound signals from the abdominal wall surface (Fig. 1a,b).

**Figure. 1.**
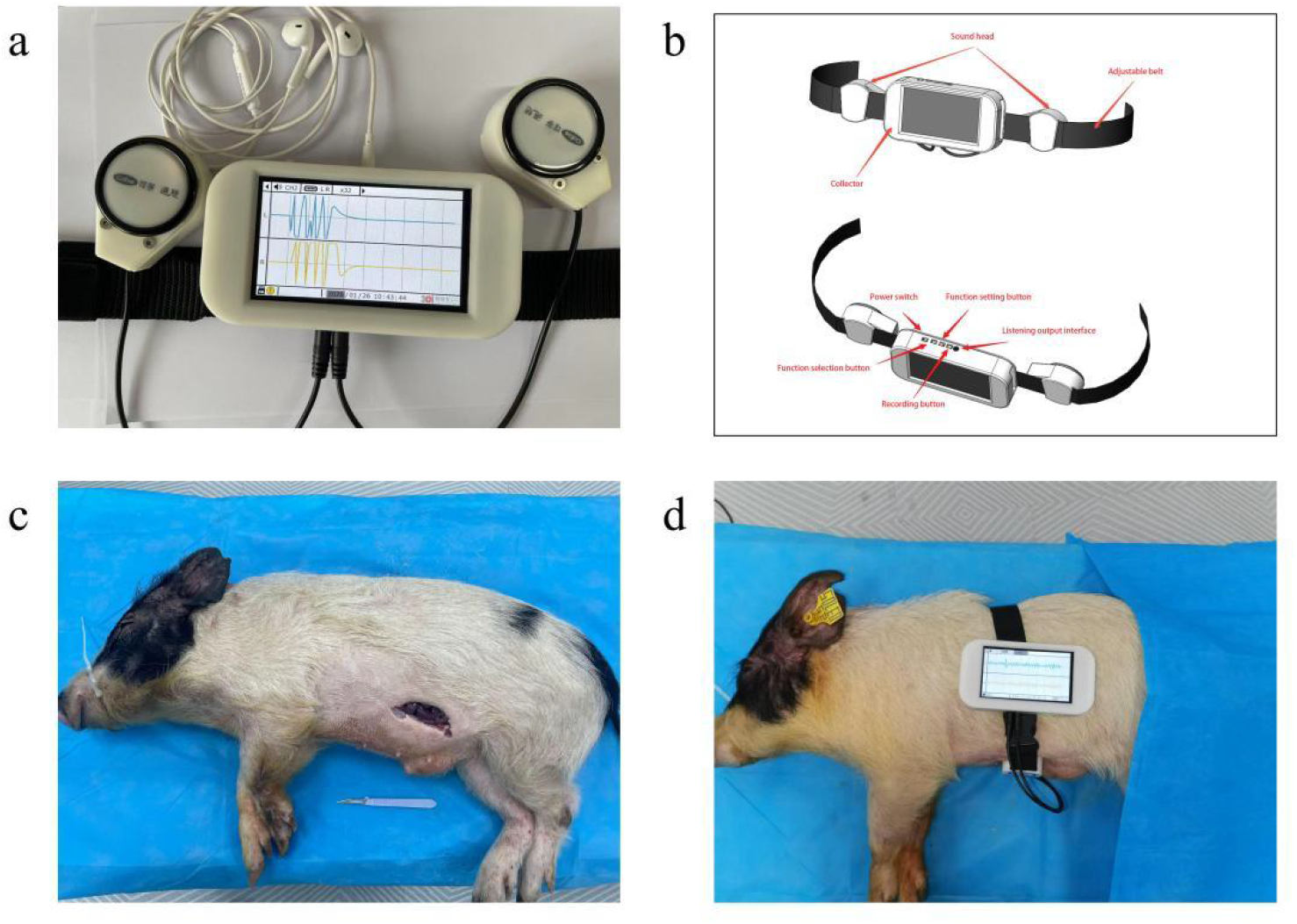
Experimental configuration of the bowel sound monitoring system. **a**, Photograph of the self-developed bowel sound monitoring device. **b**, Schematic illustration of the device function. **c**, After skin preparation, a lateral abdominal incision was made in a Bama miniature pig, and a wireless acoustic source was placed within the abdominal cavity before fixation and closure. **d**, The bowel sound monitoring device was secured to the abdominal surface for signal acquisition.

The device was designed to balance clinical usability with portability. It measures 14.6 × 7.6 × 3.3 cm and features an ergonomic form factor suitable for handheld operation. A 4.3-inch high-definition display, supporting 24-bit colour depth and a resolution of 800 × 480 pixels, enables clear visualization of audio waveforms. Audio acquisition is achieved through a dual-chestpiece, dual-channel stereo configuration, with each sensing head measuring 47 mm in diameter, making the device compatible with clinical auscultation scenarios and capable of effective bowel sound detection, continuous monitoring, and accurate interpretation.

The device is powered by an STM32F429IGT microcontroller with an ARM Cortex-M4 core operating at 180 MHz, supporting real-time audio signal processing and smooth multitask operation. For audio acquisition, the system supports sampling rates from 8 kHz to 96 kHz and sampling depths of 16, 20, 24, and 32 bits, covering the full frequency range required for medical auscultation. Combined with high-sensitivity microphones, the device enables precise capture of weak bowel sound signals. Its theoretical battery life is 33.3 h, supporting continuous monitoring across different clinical scenarios.

Overall, the device demonstrated robust capabilities in acoustic signal acquisition, processing, and display, providing a solid technical foundation for reliability validation under realistic tissue acoustic propagation conditions. The detailed configuration parameters of the self-developed bowel sound monitoring device are summarized in Table 1.

**Table 1.**
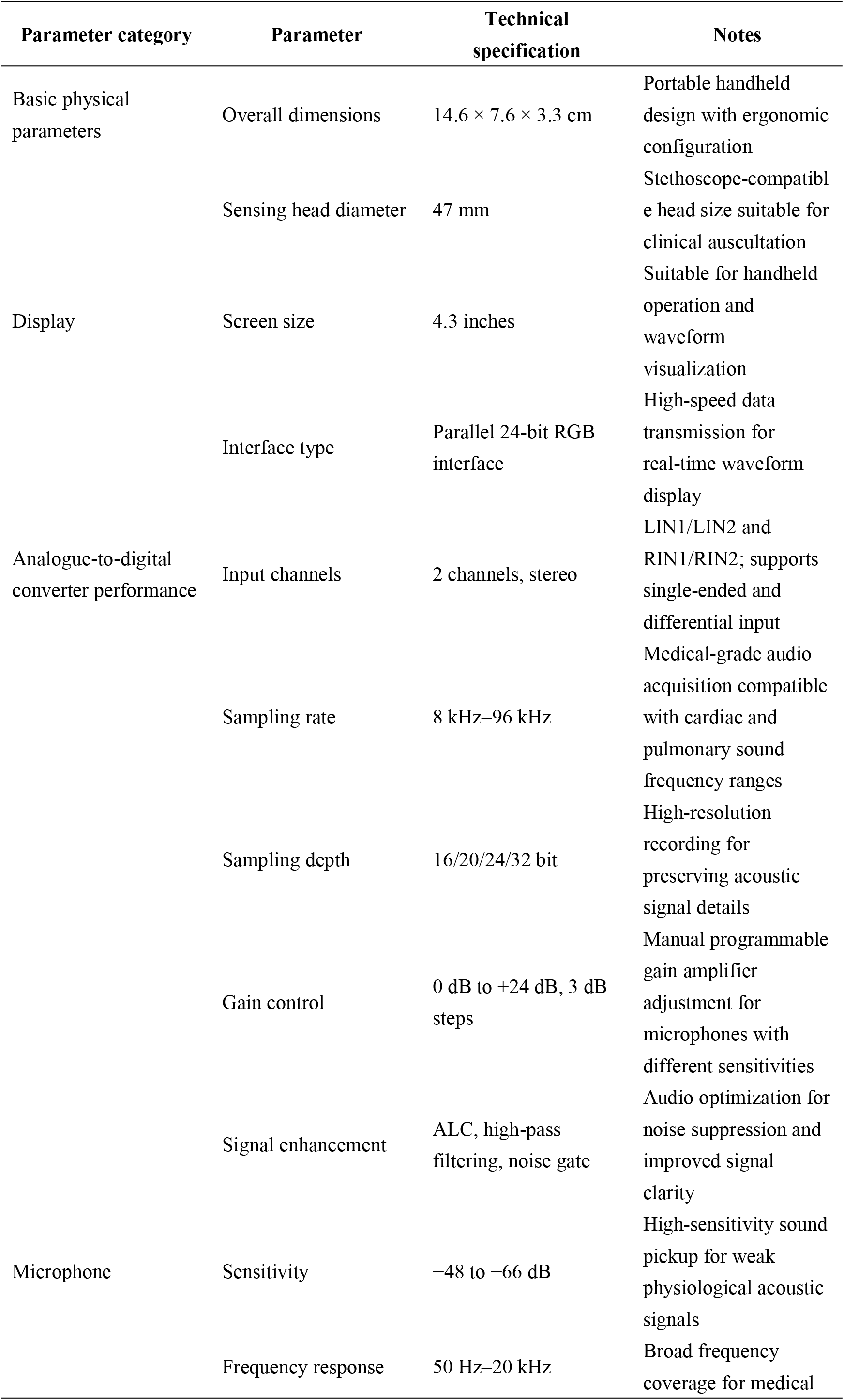
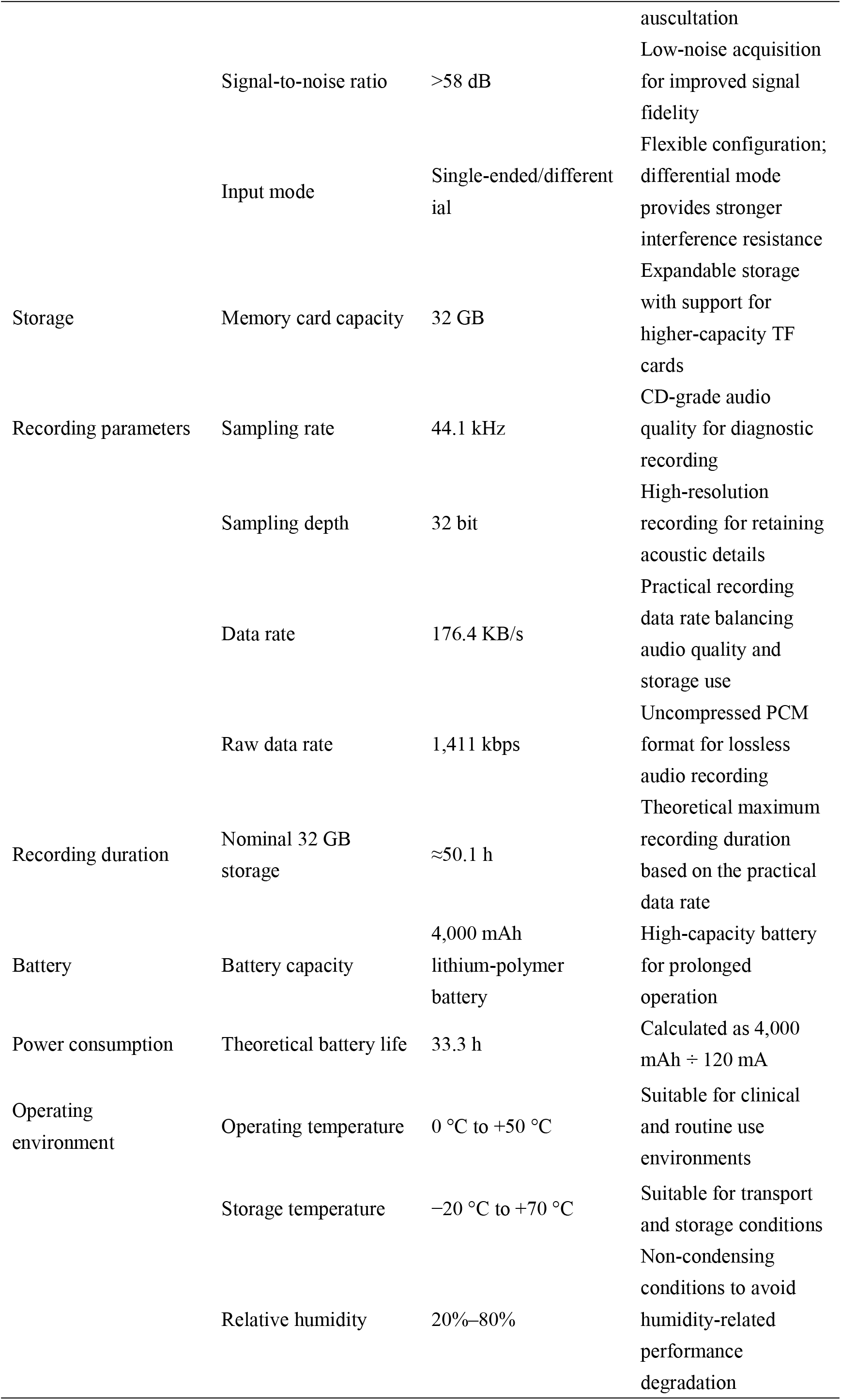

The device acquires, stores and time-stamps acoustic signals according to predefined sampling parameters, thereby supporting subsequent event-level analysis.

A high-fidelity Bluetooth audio playback device, Bose SoundLink Revolve II, USA, was used as the experimental acoustic source. The device was placed within the abdominal cavity to play manually annotated bowel sound recordings as controlled acoustic stimuli. This acoustic source was used solely to provide reproducible and controllable bowel sound stimuli and did not perform any physiological monitoring or diagnostic function.

#### 1.2.2 Preparation of bowel sound recordings and playback settings

The bowel sound recordings used in this study were obtained from a publicly available database containing manually annotated standard bowel sound data, available at https://doi.org/10.6084/m9.figshare.28595741. All recordings contained explicit timestamp annotations for bowel sound events. According to previous classifications of bowel sound patterns, the annotated events in the source recordings were categorized into four types: single burst, SB; multiple burst, MB; continuous random sound, CRS; and harmonic sound, HS [9]. The audio files were stored in standard digital audio formats and covered bowel sound events with varying durations and acoustic morphologies.

Before the experiment, the temporal sequence and inter-event intervals of the playback recordings were standardized to ensure that individual bowel sound events remained independent and to prevent overlapping events from interfering with subsequent event matching and analysis. No external power supply or wired connection was used during the experiment. All playback procedures were performed wirelessly to minimize potential mechanical interference with the intra-abdominal acoustic propagation pathway. Playback parameters were kept constant throughout the experiment.

#### 1.2.3 Experimental procedure and data acquisition

A surgical opening was created in the lateral abdominal wall of a freshly euthanized Bama miniature pig. The Bluetooth audio playback device was placed into the abdominal cavity, and the incision was closed by suturing to reduce the influence of acoustic source displacement on propagation conditions during the experiment (Fig. 1c). The self-developed bowel sound monitoring device was then fixed to the abdominal surface, ensuring stable contact with the skin.

After device placement, bowel sound playback and continuous surface audio acquisition were initiated simultaneously. The acquisition period included both bowel sound playback intervals and non-playback control intervals, allowing complete recording of body-surface acoustic signals and corresponding temporal information (Fig. 1d). After the experiment, all recorded data were exported for subsequent event matching and quantitative analysis.

#### 1.2.4 Event matching and metric calculation

The timestamps of bowel sound events played from within the abdominal cavity were used as the ground-truth reference. Within a predefined temporal tolerance window, bowel sound events detected by the monitoring device were matched at the event level to the corresponding playback events. Based on this framework, signal acquisition and energy gain, bowel sound event matching, and acoustic feature consistency were evaluated.

In addition, gastroenterologists with different levels of clinical experience were invited to perform blinded auscultation-based annotation of the recorded audio. This analysis was used to assess the degree of perceptual agreement between the device-acquired signals and the original acoustic source at the clinical auscultation level.

### 1.3 Evaluation metrics

An event-level evaluation framework was used to systematically assess the acquisition and recognition performance of the bowel sound monitoring device under realistic tissue-mediated acoustic propagation conditions. All evaluations used the annotated intra-abdominal playback events as the ground-truth reference. Event matching and statistical analyses were performed within a predefined temporal tolerance window. The main evaluation metrics were as follows.

First, bowel sound acquisition and energy gain were assessed by comparing spectral distribution and energy changes between the original and recorded signals, thereby evaluating the device’s ability to cover and amplify the original bowel sound signals.

Second, bowel sound event matching was performed using the timestamps of the original recordings as the reference. Detected events were matched within a predefined temporal window to evaluate the temporal localization accuracy and event identification consistency of the device.

Third, acoustic feature consistency was quantified using spectral structure and Mel-spectrogram representations to assess the similarity between the recorded and original audio signals in the time–frequency domain [10].

Fourth, subjective agreement in clinical auscultation was evaluated. Two attending gastroenterologists and two associate chief gastroenterologists were recruited from the Department of Gastroenterology. All had more than 5 years of specialized clinical experience in gastroenterology and extensive experience in abdominal physical examination and the diagnosis and treatment of common gastrointestinal disorders. In a blinded manner, they manually annotated randomly presented original source recordings and body-surface recordings. The annotation consistency of bowel sound events between the two types of audio was compared to evaluate the clinical auscultation-level reliability of the self-developed monitoring device [5,11–15].

Together, these metrics provided a multidimensional evaluation of device reliability, including event detectability, temporal localization accuracy, acoustic feature consistency and subjective perceptual agreement. No diagnostic or physiological functional inference was made.

### 1.4 Statistical analysis

Descriptive statistics were used to analyse the experimental data. Categorical variables are presented as counts and percentages. Continuous variables are presented as mean ± standard deviation or median with interquartile range, depending on data distribution. Event detection rate and other evaluation metrics were calculated and analysed according to predefined definitions. Data processing and statistical analyses were performed using SPSS version 22.0.

## 2. Results

### 2.1 Bowel sound acquisition and energy gain

Bowel sound monitoring was performed under realistic tissue-mediated acoustic propagation conditions. During the experiment, manually annotated standard bowel sound recordings were played continuously for 60 min as the source audio, while the self-developed bowel sound monitoring device continuously acquired body-surface audio signals as the recorded audio. The acquired data fully covered the playback period. For event-level matching and analysis, 90-s audio segments were randomly selected every 15 min. No data were excluded because of signal loss or acquisition failure.

Comparison of the mean spectra of the source audio, shown in orange, and the recorded audio, shown in blue, demonstrated that the frequency distribution of bowel sounds was mainly located within the 100–2,000 Hz range. The recorded audio covered the full spectral range of the original audio (Fig. 2a). The two types of audio showed highly similar spectral structures. The dominant energy of the recorded audio was concentrated within the 200–300 Hz range, corresponding to the main spectral peak, and the waveform pattern in this peak region was preserved. The overall spectral morphology consistency was 86.9%, and the area-under-the-curve overlap between the two spectra was 72.8%.

**Fig. 2.**
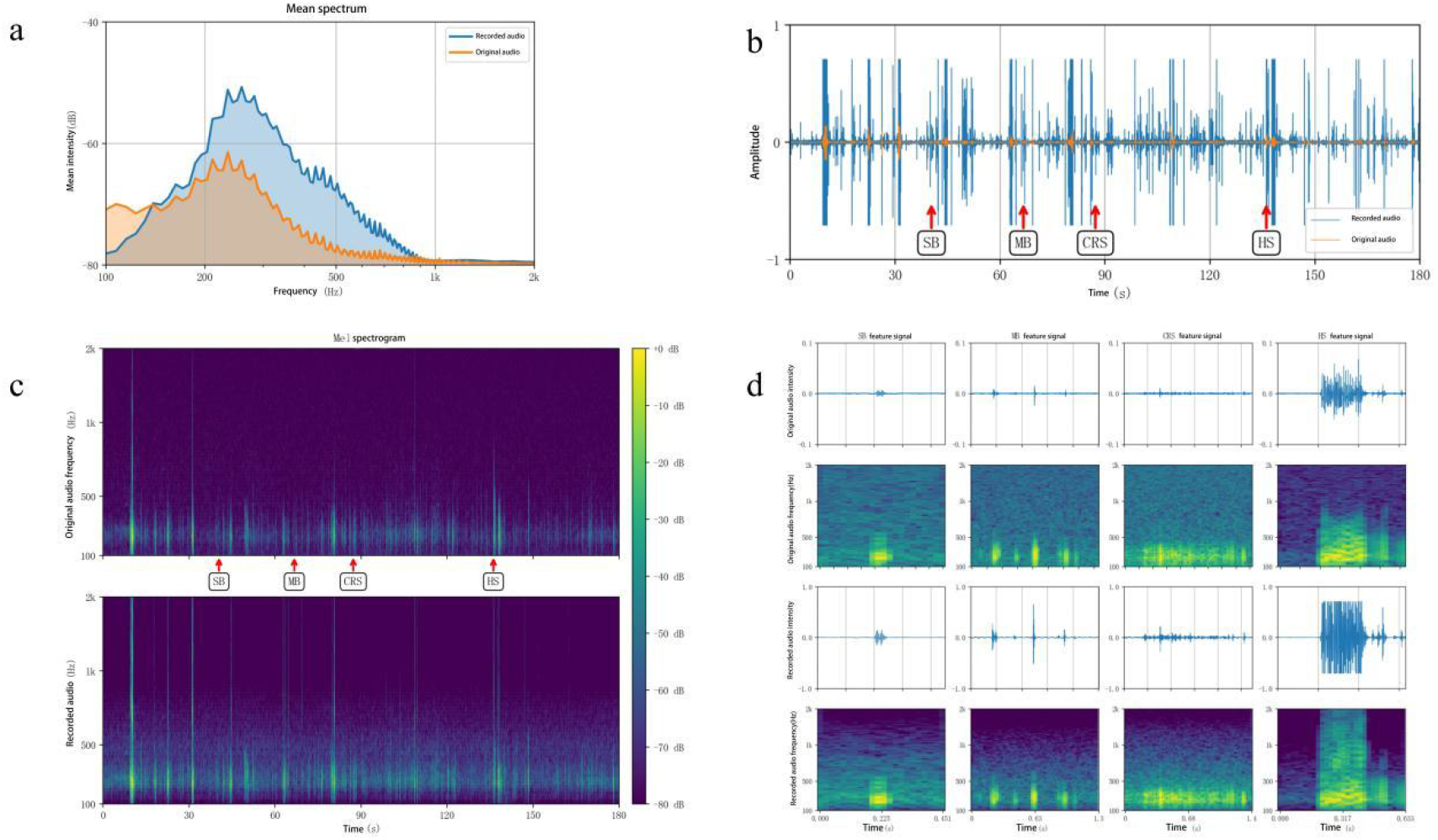
Comparative analysis of source and recorded bowel sound audio. a, Waveform comparison between the source audio and the recorded audio. The recorded audio is shown in blue and the source audio in orange. b, Mean spectral comparison between the recorded and source audio. The recorded audio is shown in blue and the source audio in orange. c, Mel-spectrograms of the source and recorded audio. d, Preservation of characteristic bowel sound patterns in the recorded audio.

These findings indicate that, under realistic biological tissue acoustic propagation conditions, the bowel sound monitoring device effectively amplified the original bowel sound signals while preserving their spectral characteristics, thereby facilitating clinical recognition of bowel sound events.

### 2.2 Bowel sound event matching

The waveform comparison between the original and recorded audio is shown in Fig. 2b. Compared with the original audio waveform, shown in orange, the recorded waveform, shown in blue, exhibited a markedly increased amplitude. Importantly, all waveform segments corresponding to changes in the original signal were effectively retained in the recorded audio. These results indicate that the device can amplify bowel sound signals while preserving their temporal-domain characteristics.

### 2.3 Acoustic feature consistency analysis

In this study, Mel spectrograms were used to represent bowel sound signals and to bridge the gap between physical frequency and physiological perception. A Mel spectrogram remaps the conventional frequency spectrum according to human auditory perception by modelling the nonlinear frequency response of the human auditory system. This representation provides higher resolution for low-frequency signals and moderate compression of high-frequency components, thereby improving the discriminability of bowel sounds in complex background noise.

As shown in Fig. 2c, the temporal distribution and frequency distribution of the highlighted segments in the source audio, shown in the upper panel, were well preserved in the recorded audio, shown in the lower panel. These findings indicate that the device effectively retained the time-domain and frequency-domain characteristics of bowel sound signals.

To further assess the preservation of characteristic bowel sound patterns in the recorded audio, four representative audio segments were extracted according to the annotations of the source audio, corresponding to the four bowel sound patterns (Fig. 2d). Within the bowel sound frequency range, the recorded audio showed highly consistent spectral characteristics with the source audio for each sound pattern, indicating that the recording method effectively preserved the time–frequency features of different bowel sound types. In addition, the recorded spectrogram showed a clear contrast between low-frequency and high-frequency regions, suggesting that the recording method may enhance the signal-to-noise ratio of bowel sounds and facilitate clearer clinical identification of bowel sound events.

### 2.4 Subjective consistency of physician auscultation

Finally, four clinicians with different levels of clinical experience were recruited to perform blinded auscultation-based annotation of the source audio and the body-surface recorded audio in a self-controlled manner. The overall annotation agreement for bowel sound events between the two types of audio was 88.9%. Agreement was 84.9% in the attending physician group and 92.9% in the associate chief physician group, with no significant difference between physicians of different seniority.

Taken together, these results demonstrate that the self-developed bowel sound monitoring device can stably detect bowel sound events under realistic tissue acoustic propagation conditions, with favourable overall consistency in temporal localization, acoustic feature fidelity and resistance to false-positive detection.

## 3. Discussion

Bowel sounds are objective acoustic signals that reflect gastrointestinal motility and have been explored for non-invasive estimation of bowel motility [16]. However, their short duration, sparse occurrence, complex acoustic morphology and susceptibility to environmental noise have made continuous, objective and standardized acquisition technically challenging in clinical practice and research [17]. In this study, we used a controlled acoustic source to systematically validate the acquisition and recognition reliability of a self-developed bowel sound monitoring device under realistic biological tissue acoustic propagation conditions. The aim was to provide an engineering and methodological basis for bowel sound research and future clinical translation.

The results showed that the device achieved high sensitivity for detecting bowel sound events under realistic tissue propagation conditions, suggesting that it can stably capture bowel sound signals along the complex abdominal cavity–abdominal wall–body surface acoustic transmission pathway. Compared with conventional approaches that rely mainly on manual auscultation or short-duration, discontinuous acquisition, this device offers clear advantages for continuous monitoring and event-level acquisition. It may therefore reduce the influence of subjective human judgement and provide a technical foundation for objective bowel sound recording [9].

A methodological strength of this study is the inclusion of blinded physician auscultation agreement as a complementary validation to objective event-level metrics [3]. In clinical practice, bowel sounds have traditionally been interpreted through physician auscultation, and their perceptibility and interpretability by the human ear remain central to their clinical relevance. By asking gastroenterologists with different levels of seniority to blindly annotate both source audio and body-surface recordings, and by comparing agreement in bowel sound event identification between the two audio types, this study evaluated the validity of the recorded signals from a clinical perceptual perspective [18].

Unlike validation approaches based solely on algorithms or engineering indices, blinded physician agreement reflects the acceptability of the device-acquired signals within the clinical auscultation context, without introducing any diagnostic inference. This combined framework, integrating objective acoustic metrics with subjective auscultation agreement, helps narrow the gap between engineering performance assessment and clinical application, and provides more interpretable evidence for the clinical translation of bowel sound monitoring devices.

It should be noted that this study used freshly euthanized Bama miniature pigs as the experimental model. This design preserved, as far as possible, the acoustic propagation characteristics of real biological tissue, while introducing manually annotated bowel sound recordings as a controlled acoustic source.This approach reduced the uncertainty associated with uncontrollable bowel sound timing and ambiguous event annotation in live-animal experiments, following the broader logic of controlled model-based qualification for acoustic monitoring sensors [19].The purpose of this study was not to simulate active intestinal physiology. Rather, by constructing a defined and reproducible real-tissue acoustic propagation environment, we focused on validating the device’s ability to acquire and identify bowel sound events. This “controlled acoustic source plus real tissue propagation” framework provides a feasible and reproducible technical pathway for engineering validation of bowel sound monitoring devices.

This study has several limitations. First, no live animals or clinical participants were included; therefore, the performance of the device under complex physiological conditions remains to be evaluated. Second, although the playback audio was derived from real annotated bowel sound data, it may not fully capture the complete acoustic spectrum of bowel sounds across different pathological states. Third, this study mainly focused on event-level detection and temporal consistency, whereas changes in signal intensity, detailed spectral characteristics and long-term monitoring stability were not comprehensively analysed.

Despite these limitations, this study validates the reliability of a self-developed bowel sound monitoring device under realistic tissue acoustic propagation conditions from an engineering perspective. These findings provide a foundation for subsequent live-animal experiments, clinical validation and algorithm optimization. Future studies should further combine live-animal models and clinical cohorts to systematically evaluate the performance of this device under different physiological and pathological conditions, and to explore its potential applications in gastrointestinal function assessment, perioperative monitoring and remote health management.

## Data Availability

All data produced in the present study are available upon reasonable request to the authors
All data produced in the present work are contained in the manuscript
All data produced are available online at

## Conflict of interest

**The authors declare no competing interests**.

## Author contributions

Jingxuan Zhao contributed to study design, overall implementation, data analysis and manuscript drafting. Zhizhuang Zhao, Xianya Huang and Yuxuan Li contributed to data acquisition and resource support. Junling Wu contributed to experimental and resource support. Shufang Wang and Gang Sun contributed to study design, manuscript review and resource management. Zhe Luan supervised the overall study design, implementation, analysis and final manuscript preparation.

## Notes

### Competing Interest Statement

The authors have declared no competing interest.

